# First Measurement: Proactive Immune Cell Sparing in Radiation Therapy

**DOI:** 10.1101/2025.01.05.25320011

**Authors:** Krishni Wijesooriya, Cam Nguyen, Mark R Conaway, Paul W Read, Kara Romano, Lawrence G Lum, Archana Thakur, David W Lain, Christopher M McLaughlin, Christopher K Luminais, Song Wood, Grant Williams, Joe Chen, Bryant Walker, Damon Sprouts, Donald Muller, Kristin Ward, Sunil Dutta, Jason Sanders, David Cousins, Ebenezer Asare, Emily Nesbit, Yhana C Chavis, Kristin V Walker, Einsley Janowski, Timothy Showalter, James M Larner

**Author notes:** **Correspondence to:** Krishni Wijesooriya, PhD, University of Virginia Department of Radiation Oncology.

## Abstract

**Purpose:** Radiation Therapy (RT) can modulate the immune system and generate anti-tumor T cells. However, this anti-tumor-activity is countered by radiation-induced immunosuppression (RIIS). Clinical advantages of proactively sparing RT dose to immune rich organs have not previously been evaluated.

**Methods:** We conducted a phase II randomized trial from 2020 to 2023, enrolling 51 early-stage lung cancer patients treated with SBRT, to evaluate the effect of dose reduction to immune rich organs on RIIS. Two groups were: RIIS-optimized-treatment (lowering the dose to blood, bone-marrow and lymph-node-stations) and standard-treatment. All treatments followed national protocol guidelines. Peripheral blood was collected at baseline, immediately, 4-weeks and 6-months post-treatment.

**Results:** ALC changes from baseline immediately, 4-weeks and 6-months post-SBRT are: optimized-arm: -16%, -22%, -16%, standard-arm: -31%, -34%, -26%, leading to an overall-all-time-point improvement in ALC reduction in the optimized-arm compared to the standard-arm of 13.4 (5.3) % (95% CI, 2.8 to 24.0; *p* = 0.01). Central tumors had the largest improvement in ALC from baseline: optimized-arm: - 8%, -18%, -14%, standard-arm: -39%, -43%, -47%, leading to an overall-all-time-point improvement in ALC reduction in the optimized-arm compared to the standard-arm of 29.5 (9.6) % (95% CI, 10.1 to 48.9; *p* = 0.004).

Grade 3 lymphopenia occurred in 15.4% of standard arm patients but was absent in the optimized arm. Additionally, 2.8 times more patients in the optimized arm experienced an ALC increase post-SBRT. Dose to organs such as the heart, great vessels, thoracic spine, and lymph nodes significantly correlated with RIIS.

A trend towards increased Event-Free-Survival (two-year: 75.0% (SE = 10.8%) versus 59.8% (SE = 11.2%), *p*=0·10) and Overall Survival (two-year: 93.4% (SE=6.1%) versus 69.4% (SE=10.5%), *p*=0·14) was observed with optimized-planning compared to standard-planning in treatment naïve patients.

**Conclusion:** Reducing RT dose to immune rich organs significantly reduces RIIS compared to standard-of-care. This has implications in enhancing immune system mediated anti-tumor-activity. (Funded by National Cancer Institute and others. ClinicalTrials.gov number, NCT04273893)

## Introduction

Lymphocytes play a crucial role in the body’s response to cancer by directly attacking tumors, inhibiting tumor growth and spread, and detecting and potentially eliminating emerging tumors (1,2). Tumor-infiltrating lymphocytes (TILs) are commonly found within tumors, and studies have shown that their presence is associated with more favorable outcomes (3-6). Radiation therapy (RT), particularly Stereotactic Body Radiation Therapy (SBRT), modulates the immune system by promoting the generation of Cytotoxic T Lymphocytes (CTLs) and enhancing T cell infiltration into tumors (7). These CTLs may eliminate distant metastases or residual disease (abscopal effect) not targeted by the primary treatment (8).

Radiation is known to induce immunosuppression (radiation-induced immunosuppression, RIIS) and lymphopenia (10-12), an unavoidable side effect of RT, which may impede tumor control, reduce survival, and increase hospitalizations (13 -27). Lymphocytes, 80% of which are T cells, are highly radiosensitive, and RIIS destroys existing as well as newly recruited CTLs.

A recent prospective study involving patients with multiple tumor types (9), including Non-Small-Cell Lung Cancer (NSCLC), receiving concurrent RT and immunotherapy demonstrated that higher post-RT absolute lymphocyte counts (ALC) correlated with improved abscopal responses, while lower post-RT ALC was associated with poorer progression-free survival (*p* =.009) and overall survival (*p* = .026).

The findings suggest that minimizing radiation exposure to immune cells could maximize the therapeutic ratio and improve clinical outcomes. Thoracic tumors are adjacent to many blood/immune rich organs including the great vessels, heart, thoracic-spine, and lymph-node-stations. As a result, the impact of RT to lymphocytes can be significant. Current RT treatment planning guidelines do not account for RIIS. Optimizing RT to minimize RIIS by considering circulating blood or lymphatics as critical Organs At Risk (OAR) is not supported by existing algorithms.

We present the results of a phase II randomized clinical trial (**NCT04273893**) (28) investigating the proactive reduction of radiation dose to blood- and lymph-rich organs that mitigate RIIS during lung SBRT, compared to the standard of care.

## Methods

### Trial Design

This trial compared two radiation therapy planning techniques for early-stage lung cancer: standard of care SBRT planning that meet Radiation Therapy Oncology Group (RTOG) 0813/0915 (29,30) dosimetric criteria and optimized SBRT planning that meet standard of care dosimetric criteria while minimizing radiation exposure to circulating blood and lymphatics, including the heart, great-vessels, lymph-node-stations, and thoracic-spine.

The protocol (IRB #21718), was approved by the Institutional Review Board and the ethics committee, monitored by the institutional data and safety monitoring committee, and conducted in accordance with the Declaration of Helsinki and Good Clinical Practice. All patients provided written informed consent. The trial had interim toxicity analysis.

### Patients

We planned to enroll 50 patients (25 in each arm) who were 18 years or older, had an Eastern Cooperative Oncology Group (ECOG) performance status of 0–2, demonstrated adequate organ function, and had pathologically or imaging-confirmed early-stage NSCLC according to the American Joint Committee on Cancer version 8 staging system. Eligible patients were those unable or unwilling to undergo surgery. Patients with recurrence of previously treated NSCLC were also eligible, provided no further surgery was planned and they met the other eligibility criteria.

All patients had pre-radiation therapy ALCs > 0.5 × 10^9^ cells/L (Grade 3 lymphopenia) (31), within two weeks prior to registration.

Exclusion criteria included: a prior history of radiation therapy within the past two years, systemic anti-cancer therapy within the year prior to registration or planned during or within six months post-SBRT follow-up period, and major surgery within 30 days before registration or planned before the completion of the six-month post-SBRT.

### Radiation therapy treatment planning

All participants underwent four-dimensional CT simulation-based motion management with Body Fix immobilization and an Aquaplast mask for upper lobe tumors near the apex. An internal target volume (ITV) was created from 10 respiratory phases for tumors with <1 cm motion, while motion management (abdominal compression, respiratory gating) was used otherwise. A planning target volume (PTV) margin of 5mm radially and 8mm superiorly/inferiorly was added to the ITV. Treatment plans followed RTOG 0813 and 0915 guidelines, delivering SBRT in 5 fractions (45-60 Gy) via intensity modulated radiation therapy (IMRT) or volumetric modulated arc radiotherapy (VMAT), with a chest wall constraint of 30cc receiving less than 30 Gy. Varian Trilogy or TrueBeam systems were used with daily pre-treatment cone beam CT for tumor alignment.

All organs (except lungs) were manually contoured and reviewed following the RTOG atlas for organs at risk in thoracic radiation therapy, along with lymph-node-stations as defined in Chapet et al. (32), encompassing a region of interest up to a low threshold dose of 40 cGy per fraction; it has been reported that 50 cGy is a threshold for lymphocyte kill (33). All radiation therapy treatment planning were performed in Pinnacle treatment planning system, using 6X-FFF beams (except one patient with 6X beams).

### Follow-up visits

Follow-up visits, including clinical exams and CT or PET-CT imaging, were scheduled per standard of care every 3-6 months, for 3 years, and annually thereafter. Suspected recurrent disease findings were assessed by a multidisciplinary team unaware of treatment assignments.

An “event” encompassed local recurrence (in-field regrowth or new disease anywhere in the same lobe), regional recurrence (any intrathoracic lymph node), distant metastasis (all extra-thoracic areas, along with lung disease in any separate lobe), second primary lung cancer, or death from any cause.

### Quantitative analysis of blood counts

Serum and peripheral blood were collected at baseline, end-of-treatment, four-weeks and six-months post treatment. Blood tests were performed by health system’s medical lab. Absolute neutrophil, hemoglobin, platelet, and lymphocyte counts were measured, and changes between groups were compared. ALCs were also compared to simulation predictions and a multivariate analysis to explore correlations between ALC reduction, radiation dose volumes to blood- and immune-rich organs.

### Endpoints

The primary endpoints were to i) determine whether lymphocyte-sparing SBRT planning (Arm 1: investigational) results in less *in vivo* lymphocyte depletion compared to standard SBRT planning (Arm 2: control) at end-of-treatment, 4-weeks, and 6-months post treatment, ii) compare the safety and toxicity between the two arms, including the frequency, severity, and duration of grade III or IV adverse events (based on CTCAE 5.0 criteria), and iii) assess the performance of an algorithm that predicts post-treatment lymphocyte count reduction (presented in a separate manuscript).

### Statistical Analysis

Participants were randomly assigned (1:1) to Arm 1 or Arm 2 using the Clinical Trial Conduct website, stratified by tumor location (central or peripheral), using permuted blocks of sizes 2 and 4. Treatment assignments were not masked.

Analyses of absolute and percentage changes from baseline in ALC and other blood cells were conducted using repeated measures models with an unstructured covariance matrix and model terms for tumor location, follow-up time, group, and group-by-time interaction. Adjustments for stratification factors were made using least squares means. F-tests based on contrasts with the Kenward-Roger approximation to degrees of freedom were used for specific time-point comparisons, and two-sample t-tests with unequal variances were used to compare dosimetric parameters between groups. A *p* value less than 0.05 was considered significant. Descriptive statistics summarized other endpoints. Three unplanned subgroup analyses: i) Kaplan-Meier estimates and log-rank tests, evaluated the impact of dose reductions on Event-Free-Survival (EFS) and Overall-Survival (OS) ii) repeated measures analyzed relationships between clinical factors (e.g., Age, Sex, location) and their interaction between treatment and the effect on ALC reduction iii) Spearman’s correlation analyzed relationships between dose volumes to immune-rich organs and their effects on ALC reduction. Post hoc analyses were exploratory.

For all participants and within combinations of tumor location and tumor volume, interaction terms were used to test whether the effect of the optimized plan versus the standard plan differed by assessment time. When the interaction term was not significant, an overall effect of the optimized plan versus the standard was estimated from the model that did not include the interaction terms.

## RESULTS

### Patients

Between February 12, 2020, and April 5, 2023, a total of 55 patients were enrolled in the trial and underwent randomization. Four patients withdrew or did not meet the study criteria (Figure 1).

**Figure 1.**
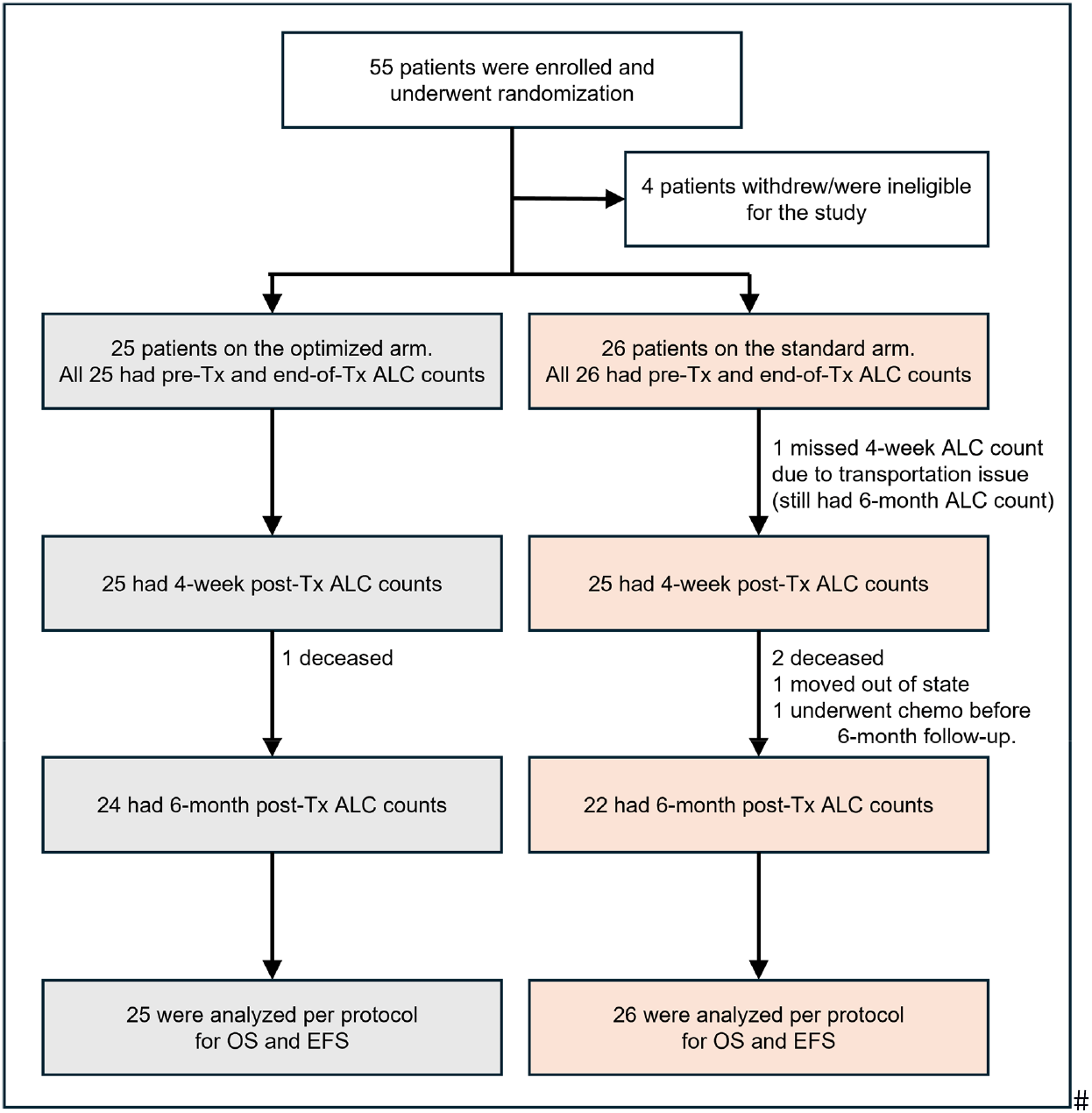
Randomization, Treatment, and Follow-up.

Number of patients (optimized; standard) completed blood draws at each time point were: end-of-treatment (25;26), 4-weeks post-treatment (25;25), 6-months post-treatment (24; 22). The database lock took place on June 30^th^ 2024, for the final analysis.

Table I provides baseline characteristics. Pre-treatment ALC results ranged from 0.79×10^9^ and 2.97×10^9^ cells/L. Despite randomization, there are some differences between the two arms including a greater percentage of patients with central tumors in the optimized arm (36.0% vs 23.1%); 3 patients in the standard arm had tumor diameter >3 to ≤5 cm while none in the optimized arm, and a higher number of treatment naïve patients in the standard arm (88.5% vs 64.0%).

**Table I:** Baseline Characteristics of the Patients.

| Characteristic | Optimized Arm | Standard Arm |
| --- | --- | --- |
| Number of patients | 25 | 26 |
| Sex |  |  |
| Male | 14 (56%) | 10 (38.5%) |
| Female | 11 (44%) | 16 (61.5%) |
| Race |  |  |
| White | 22 (88%) | 25 (96.2%) |
| Any other race | 3 (12%) | 1 (3.8%) |
| Age |  |  |
| Median (range) (years) | 74 (56-90) | 74 (55-89) |
| Smoking status |  |  |
| Current | 7 (28%) | 9 (34.6%) |
| Ex-smoker | 15 (60%) | 15 (57.7%) |
| Life-long nonsmoker | 3 (12%) | 2 (7.7%) |
| Eastern Cooperative Oncology Group (ECOG) performance status score |  |  |
| 0 | 7 (28.0%) | 3 (11.5%) |
| 1 | 15 (60.0%) | 20 (76.9%) |
| 2 | 3 (12.0%) | 3 (11.5%) |
| Tumor location |  |  |
| Central | 9 (36.0%) | 6 (23.1%) |
| Peripheral | 16 (64.0%) | 20 (76.9%) |
| Tumor histology |  |  |
| Squamous carcinoma | 2 (8.0%) | 3 (11.5%) |
| Adenocarcinoma | 7 (28.0%) | 8 (30.8%) |
| Non-small cell not otherwise specified | 16 (64.0%) | 15 (57.7%) |
| Tumor size |  |  |
| Median (range) (cm) | 1.50 (0.80 - 2.70) | 1.55 (0.90 - 4.50) |
| ≤2 cm | 18 (72.0%) | 15 (57.7%) |
| >2 to ≤3 cm | 7 (28.0%) | 8 (30.8%) |
| >3 to ≤5 cm | 0 (0.0%) | 3 (11.5%) |
| Pre-Tx ALC |  |  |
| Median (range) (10<sup>9</sup> cells/L) | 1.75 (1.05 - 2.59) | 1.87 (0.79 - 2.97) |
| Positron emission tomography staging |  |  |
| yes | 23 (92.0%) | 25 (96.2%) |
| No | 2 (8.0%) | 1 (3.8%) |
| Lung cancer history |  |  |
| Newly diagnosed | 16 (64.0%) | 23 (88.5%) |
| Recurrent | 9 (36.0%) | 3 (11.5%) |
| SBRT regimen |  |  |
| 45 Gy in five fractions (BED = 85.5 Gy) | 1 (4.0%) | 0 (0.0%) |
| 50 Gy in five fractions (BED = 100 Gy) | 8 (32.0%) | 8 (30.8%) |
| 55 Gy in five fractions (BED = 115.2 Gy) | 3 (12.0%) | 4 (15.4%) |
| 60 Gy in five fractions (BED = 132 Gy) | 13 (52.0%) | 14 (53.8%) |
| PD-L1 status |  |  |
| <1% | 3 (12.0%) | 3 (11.5%) |
| ≥1% | 4 (16.0%) | 6 (23.1%) |
| Unknown | 18 (72.0%) | 17 (65.4%) |
| Epidermal growth factor receptor status |  |  |
| Wild type | 6 (24.0%) | 5 (19.2%) |
| Mutated | 0 (0.0%) | 0 (0.0%) |
| Unknown | 19 (76.0%) | 21 (80.8%) |
| Clinical T stage |  |  |
| cT1 | 25 (100.0%) | 23 (88.5%) |
| cT2 | 0 (0.0%) | 3 (11.5%) |
| Clinical N stage |  |  |
| Unknown | 3 (12.0%) | 4 (15.4%) |
| cN0 | 22 (88.0%) | 22 (84.6%) |

### Radiation therapy dosimetry

A wide range of dose-volume histogram (DVH) parameters (Mean dose, integral dose: mean dose (in Gy) times the volume of the tissue (in cm3), V1 (volume receiving > 1 Gy) - V50) for the above-mentioned organs were automatically calculated and extracted from the exported DICOM treatment plan, CT data set, structure set, and the dose maps using an in-house algorithm (supplementary table I). Average percentage reductions on integral dose, V5 and V10 of optimized compared to standard plans are: aorta: 35%, 48%, 69%, heart: 21%, 43%, 68%, vena cava: 37%, 58%, 75%, thoracic-spine: 57%, 87%, 92%, lymph-node-stations: 37%, 58%, 68%, total lung-PTV: 5%, 8%, 0%, External-PTV: 16%, 20%, 19%. Figure. 2 shows five example patients comparing standard arm to optimized arm plans where tumors are located close to multiple blood and lymphatic rich organs. All five cases, 70% or more of the ITV (cyan in DVH plot) is receiving a total dose of 70 -80Gy. Patients 1, 2, and 3 are in the standard arm and patients 4 and 5 are in the optimized arm. Bottom table provides the immune rich organ doses and ALC decrease from pre-treatment value at 4-weeks post-RT. Patient 4 (PTV volume 28.9cc, largest) and patient 5 show minimal ALC reductions, while patients 1-3 show a 30-48% reduction illustrating multiple RIIS specific OAR competing for dose reduction.

**Figure 2:**
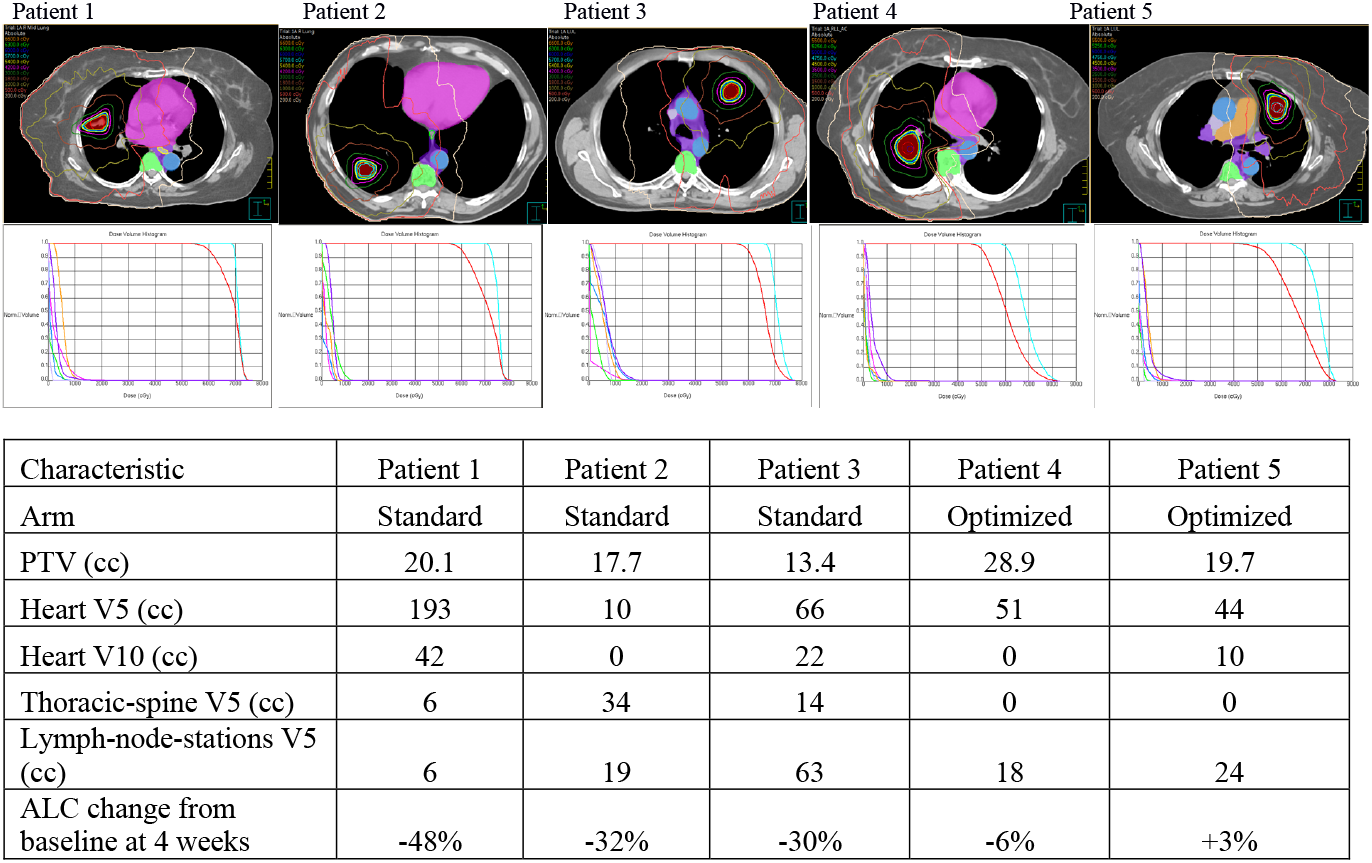
Dose distributions(top), DVHs (middle), and dose volume values for Immune rich organs (bottom) for five tumors close to heart/aorta/vena cava /thoracic-spine/lymph-node-stations. Three left hand patients had standard plans (patients 1-3), two right hand patients had optimized plans (patient 4 & patient 5). PTV (Planning Target Volume), Vx -the volume (cc) receiving a given dose x (Gy).

### Primary End Point

Supplementary Table II shows raw ALC means, unadjusted for location, for both arms at all time points. Figure 3 illustrates ALC changes from baseline for both arms, adjusted for tumor location, over time from the statistical model estimates. Left plots show absolute changes (cells/L), while right plots show percent changes from baseline. Supplementary Table III provides the ALC summaries from the statistical model estimates. Statistical-model-estimated absolute (percentage) changes from baseline ALC in the optimized arm are: 0.31×10^9^ (−16%), -0.45×10^9^ (−22%), and -0. 30×10^9^ (−16%) immediately, 4-weeks and 6-months post SBRT, respectively. Corresponding estimates in the standard arm are -0.65 x10^9^ (−31%), -0.73×10^9^ (−34%) and -0.56×10^9^ (−26%), respectively. Statistically significant between-group percent differences of 15.1% (95% CI: 3.7%, 26.5%, *p* = 0.01) immediately, 12.3% (95% CI: 0.2%, 24.5%, *p* = 0.05) 4-weeks and 10.4% (95% CI: -4.7%, 25.5%, *p* = 0.17) 6-months post SBRT were observed. Overall all time point improvement in ALC reduction (Supplementary Table IV) of 13.4% (5.3%), (95% confidence interval [CI], 2.8% to 24.0%; *p* = 0.014) in the optimized arm. Benefit of the optimized planning depends on tumor location and size (Supplementary Fig II A). Central tumors had the largest differences between the arms (Figure 3-middle plot: n=15). For central tumors, statistical-model-estimated absolute (percentage) ALC changes from baseline were -0.88×10^9^ (−39%), - 0.89×10^9^ (−43%), and -0.99×10^9^ (−50%) with the standard arm and only -0.20×10^9^ (−8%), -0.38×10^9^ (− 18%) and -0.32×10^9^ (−14%) with the optimized arm (Figure 3 middle plot), leading to an overall all time point improvement in ALC reduction of 29.5% (9.6%), (95% CI, 10.1%, 48.9%; *p* = 0.004) in the optimized arm.

**Figure 3:**
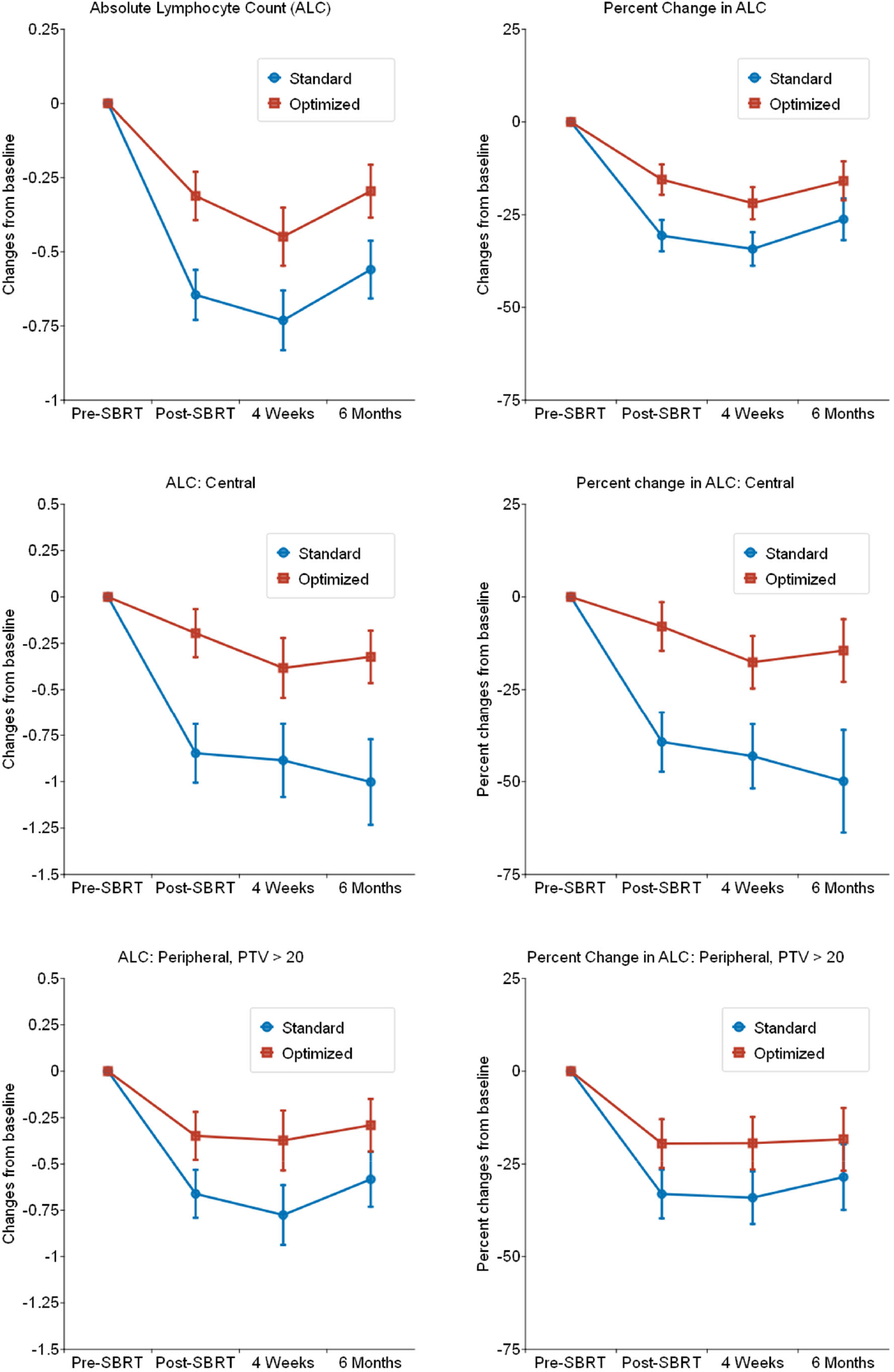
Average post-treatment lymphocyte count adjusted for stratification variable, tumor location in standard and optimized arms by assessment time. Left – average absolute ALC changes (cells/L) from baseline. Right -Percent ALC changes (%) as a fraction of baseline. Top row – all patients, middle row-central tumors, bottom row-peripheral tumors with PTV>20 cc. Optimized is the experimental arm with SBRT and standard is the standard arm with SBRT.

Peripheral tumors with PTV >20cc optimized arm displayed ameliorated immune suppression (Figure 3 bottom plot: n=18), while peripheral tumors with PTV <20cc showed no difference in ALC measurements between the two arms (Supplementary Figure I: n=18). For peripheral tumors with PTV>20cc, optimized arm observed a statistical-model-estimated overall all time point improvement in ALC reduction of 13.5% (8.6%), 95% CI, -3.8, 20.9; *p* = 0.124 compared to the standard arm.

In exploratory analyses, in the standard arm, 11.5% (all in the peripheral PTV <20cc group) of the patients had a post SBRT ALC increase at least at one time point, compared to a 32% increase (in all three sub-groups) in the optimized arm (risk difference = 20.5%, 95% CI: -1.5, 42.5%; *p* = 0.08). In the standard arm, 15.4% of patients experienced post-SBRT grade 3 lymphopenia, while none in the optimized arm were lymphopenic (risk difference -15.4%, 95% CI: -29.2%, -1.5%; *p* = 0.04). Radiation induced suppression of all blood cell types are also reduced in the optimized arm with respect to standard arm although insufficient data to determine the significance (Supplementary Figure II B).

#

### Secondary End Point

Optimized planning was associated with two grade 3 or higher adverse events, related to lung infection, while the standard arm had four grade 3 toxicities related to dyspnea, hypoxia, and lung infection (Table II). Six participants (24%) in the optimized group had grade 2 events, while nine participants (35%) in the standard group had grade 2 events. All participants recovered from their grade 3 toxicities. Grade 2 or higher pneumonitis or grade 4 or higher toxicities were not reported in either group.

**Table II:** Grade 2 or higher adverse events possibly, probably, or definitively related to therapy.

| Adverse Event | Grade 2 |  | Grade 3 |  |
| --- | --- | --- | --- | --- |
|  | Optimized Arm | Standard Arm | Optimized Arm | Standard Arm |
| Anorexia |  |  |  |  |
| Atelectasis |  | 1 |  |  |
| Bronchiolectasis |  |  |  |  |
| Cough |  |  |  |  |
| Dyspnea | 3 | 2 |  | 2 |
| Fatigue |  | 2 |  |  |
| Fracture | 1 |  |  |  |
| Generalized muscle weakness |  |  |  |  |
| Hoarseness |  |  |  |  |
| Hypoxia |  | 2 |  | 1 |
| Inflammatory changes in lung |  |  |  |  |
| Lung infection |  |  | 2 | 1 |
| Non-cardiac chest pain |  | 1 |  |  |
| Pleural effusion | 1 |  |  |  |
| Pneumonitis |  |  |  |  |
| Productive cough | 1 | 1 |  |  |
| Pulmonary fibrosis |  |  |  |  |
| Wheezing |  |  |  |  |
| Total | 6 | 9 | 2 | 4 |

### Overall-Survival (OS) and Event-Free-Survival (EFS)

Although the clinical trial was not powered for differences in survival, we evaluated the OS and EFS in order to descriptively report the outcomes of both arms (Figure 4). These numbers show a non-statistically significant trend towards improved outcomes for the optimized arm patients in both OS (*p* = 0.23) and EFS (*p* = 0.27), with two-year OS and EFS of 65.1% (SE=10.1%) and 56.7% (SE=10.5%) in the standard arm vs 82.1% (SE = 8.3%) and 62.3% (SE=10.1%) in the optimized arm. Looking specifically at treatment-naïve patients, there was a trend towards statistically significant improvement in two-year EFS of 59.8% (SE = 11.2%) in the standard arm versus 75.0% (SE = 10.8%) in the optimized arm, *p*=0·10, as well as in OS with two-year OS of 69.4% (SE=10.5%) in the standard arm versus 93.4% (SE=6.1%) in the optimized arm, *p*=0·14.

**Figure 4:**
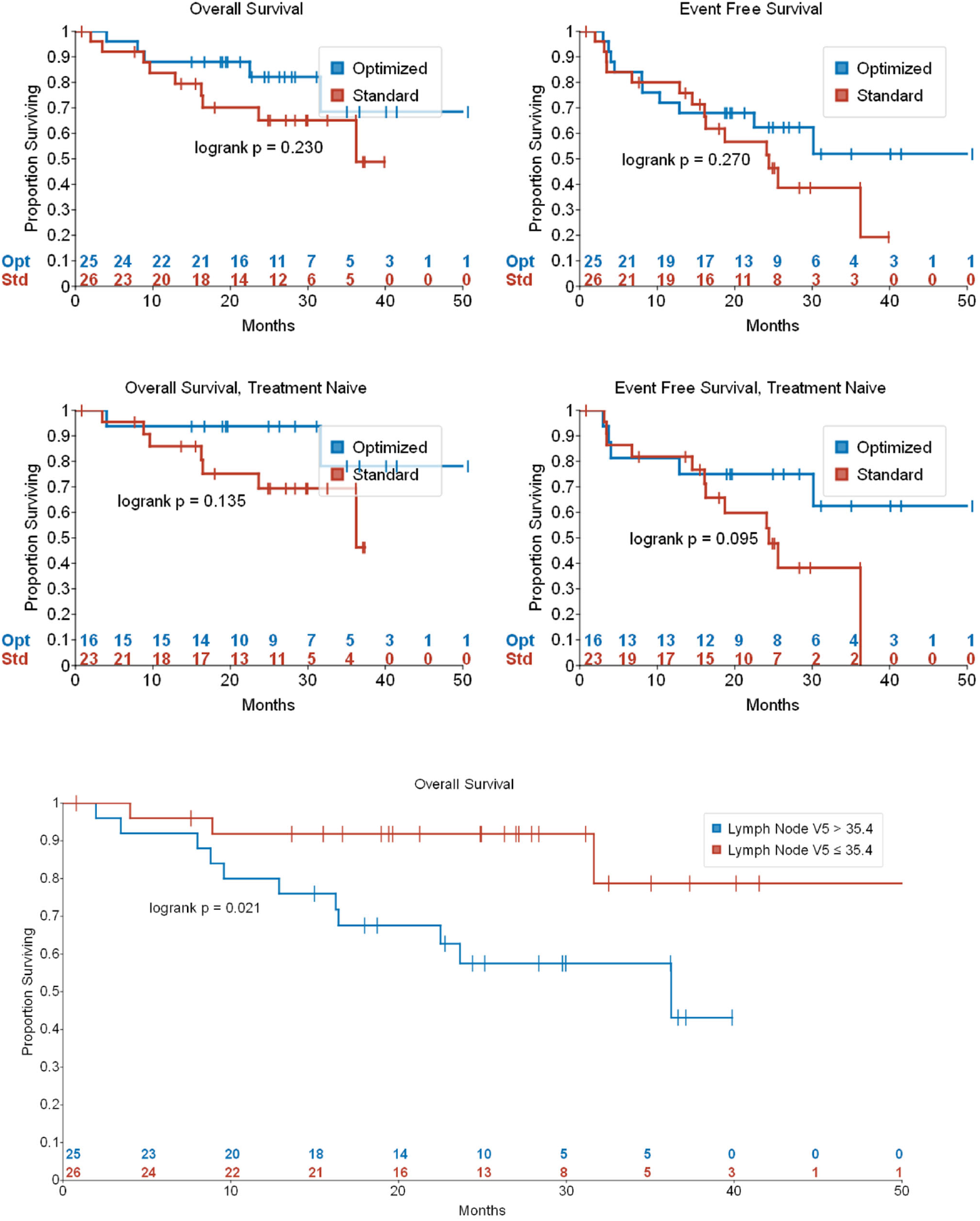
Kaplan-Meier plots of event-free survival (top left) for all patients, Overall survival (top right) for all patients, Event-free survival (middle left), and overall survival (middle right) for the treatment naïve population (n=37). E/N=events/number of participants. Optimized is the experimental arm with SBRT and standard is the standard arm with SBRT. Bottom: Kaplan-Meier plot of overall survival for all patients stratified by the volume of lymph-node-stations receiving a dose of 5 Gy by 35.4 cc (median).

### Non-dosimetric correlations on ALC drop

Demographic factors were assessed for their influence on ALC changes. Significantly higher immune sparing following optimized planning than standard planning was observed, particularly for participants who were aged < 73 years (0.54 (0.23,0.86); *p* = 0.001) (median age), female sex (0.56 (0.27, 0.86); *p*<0.001), central tumors (0.65 (0.24, 1.06); *p*=0.003), SUV maximum >3.5 (0.58 (0.26, 0.91); *p* <0.001), ECOG score of 1–2 (0.36 (0.11, 0.60) *p* =0.005), with COPD (0.36 (0.11, 0.61); *p*=0.006), and treatment naïve group (0.31 (0.05,0.57); *p*= 0.02), (Supplementary Figure III A).

### Organ-specific dosimetric correlations on ALC drop and OS

Correlation of static organ dosimetry to end of treatment changes in ALC is given in Supplementary Figure IIIB. From the immune rich organs, blood-rich organs, heart and the great vessels and lymph rich organs thoracic-spine, and lymph-node-stations V5, V10 show statistically significant correlations (*p* <0.05) with the immune suppression at end of treatment.

We tried to identify the static organ doses that significantly correlate with OS. While the number of patients who received a dose >10Gy to the heart is small due to the low number of central patients in the standard arm, there was a substantial number of patients who received a dose > 5Gy to lymph-node-stations. We see a significant (*p* =0.02) OS improvement (Figure 4, bottom plot) in the low dose cohort when we stratify the patients by the median (35.4 cc) volume of lymph-node-stations receiving a dose of 5 Gy. Two-year OS proportions are: 91.8% (SE = 5.5%) for LN V5 < 35.4 and 57.5% (SE=10.4%) for LN V5 ≥ 35.4.

## Discussion

This study provides the first clinical evidence that proactively reducing radiation dose to blood and lymphatic rich organs can significantly reduce RIIS from SBRT to lung at all time points, meeting our primary endpoint. Interestingly, 15% of the standard arm patients ended up having G3 lymphopenia while none in the optimized arm were G3 lymphopenic. Evaluations of tumor specific factors indicate a larger impact for optimized planning in central (29.5% improvement compared to standard) and larger than 20 cc PTV tumors. The trial did not see optimized-arm leading to higher adverse events. Our study indicates that optimization does not sacrifice treatment outcomes, but instead may improve outcomes, with OS and EFS trending to improvement in the optimized arm in the Tx naïve group, two-year OS (94.1% (SE=5.7%) versus 69.4% (SE=10.5%)).

The premise behind our study is that reductions in immune suppression as a part of intelligent treatment planning can yield a net immune stimulation while maintaining the ablative treatment paradigm associated with SBRT. Numerous authors, such as Schaue et al. (34), Lugade et al. (35), and Reitz et al. (36) mouse model data have demonstrated that tumor control and the number of tumor-reactive T cells increases at ablative doses, suggesting radiation-induced immune modulation. Our work indicates that the immune increase phenomenon randomly observed by some after RT is not random, and could be enabled to occur in all three patient cohorts (central, peripheral, and PTV>20cc) by carefully crafted RT dose distributions during treatment planning. Observed improvements in the immune sparing was enhanced for female sex, younger patients (<73 yrs), primary tumor SUVmax >3.5, treatment naïve status, ECOG score 1-2, BMI >25 and with COPD, some of whom are the poorest performers in general. These in fact could be used to create patient specific immune signatures. A recent study published on lung SBRT with immunotherapy showed that SBRT + immunotherapy significantly improved 4-year EFS from 53% with SBRT alone to 77% with SBRT + immunotherapy (37). Interestingly, this study showed a correlation of longer EFS with immunotherapy following SBRT and younger age, male sex, and the treatment naïve status suggesting a dependence of the immune system with these variables. These trends are of interest and deserve further validation. Immunotherapy relies on the presence of neo-antigen specific cytotoxic T cells in the body. Our work suggests that RT alone, if optimized to reduce RIIS, has the potential to improve OS and EFS - a multi-institutional randomized study that is powered to evaluate OS and EFS is needed to confirm these results.

Comprehending organ-specific dose volumes that cause RIIS during SBRT will aid in constructing predictive models for better treatment planning. With this in mind, we present the effects of organ doses on post-RT RIIS. Some notable correlations are: thoracic-spine (V5, V10, integral and mean dose), Heart +GV (V5, V10), and lymph-node-stations (V5, V10, integral and mean doses) of which are all significantly correlated with end-of-Tx RIIS. This analysis suggests that the high flux of blood flow through the great vessels and heart allows for the irradiation of a large number of lymphocytes down to a low dose area. High density of immune cells in slow moving lymphatic vessels and thoracic-spine also play a significant role in RIIS. Though statistically significant, correlation coefficients are small since the static organ doses are surrogates to moving immune cells during a dynamic RT treatment. Unlike static organs, dose to constantly moving immune cells can only be determined by a predictive algorithm that mimics the dynamic interplay between immune cell motion and dynamic dose. Since multiple RIIS specific OAR are competing for dose reduction (example in Fig 2), a patient/plan specific cumulative immune cell kill from all organs is key, instead of individual OAR-based dose limits. These plans were guided by a time dependent predictive model of RIIS that include blood flow, lymphatics flow, and osmosis of immune cells between blood and lymphatics and lymphoid organs, built by our group using retrospective data (38, 39); the comparisons will be published separately. Due to the limited number of centrally located tumors, we could not evaluate the significance of heart dose in OS. However, the analysis of lymph node stations receiving a 5Gy volume of 35.4 cc (median) was significantly correlated with OS (Two-year OS of 91.8% versus and 57.5% when stratified by LN-V5 of 35.4 cc; *p* =0.02) is no surprise given the fact that lymphocytes are the prevailing cell type in lymph nodes, constituting nearly 85% of the total immune cells. Furthermore, T cells, B cells, and dendritic cells in the human body are primarily located in the lymphatic system, which are the cells responsible for clonal expansion and adaptive immunity (40).

This study had a few limitations. First, the sample size was not adequate to give statistical power to demonstrate OS and EFS benefits. In addition, there was a low number of central tumor patients that hindered us from evaluating survival benefit on sparing blood rich organs.

Radiotherapy has been shown to induce many other immunosuppressive mechanisms in the tumor microenvironment, such as induction of Tregs (35), activation of TGFβ (41), up-regulation of PD-L1(42), induction of pro-tumorigenic M2 macrophages (43), MerTK upregulation on macrophages (44) and myeloid-derived suppressor cells (35). To alleviate some of these increases one could additionally provide anti-PD-L1 and Anti-CTLA4.

In summary, this randomized phase II trial showed that patients with early-stage lung cancer treated with SBRT have significantly reduced RIIS with optimized treatment planning. Further, this SBRT optimization correlated with decreased lymphopenia and a trend towards improved EFS and OS. A larger multi-institutional randomized trial is required to confirm these exciting results, to elucidate the specific roles of blood-rich and lymphatic-rich organs and facilitate translation into clinical practice.

## Data Availability

All data produced in the present study are available upon reasonable request to the authors

