## Supplementary material for "First Measurement: Proactive Immune Cell Sparing in Radiation Therapy": Figures and Tables

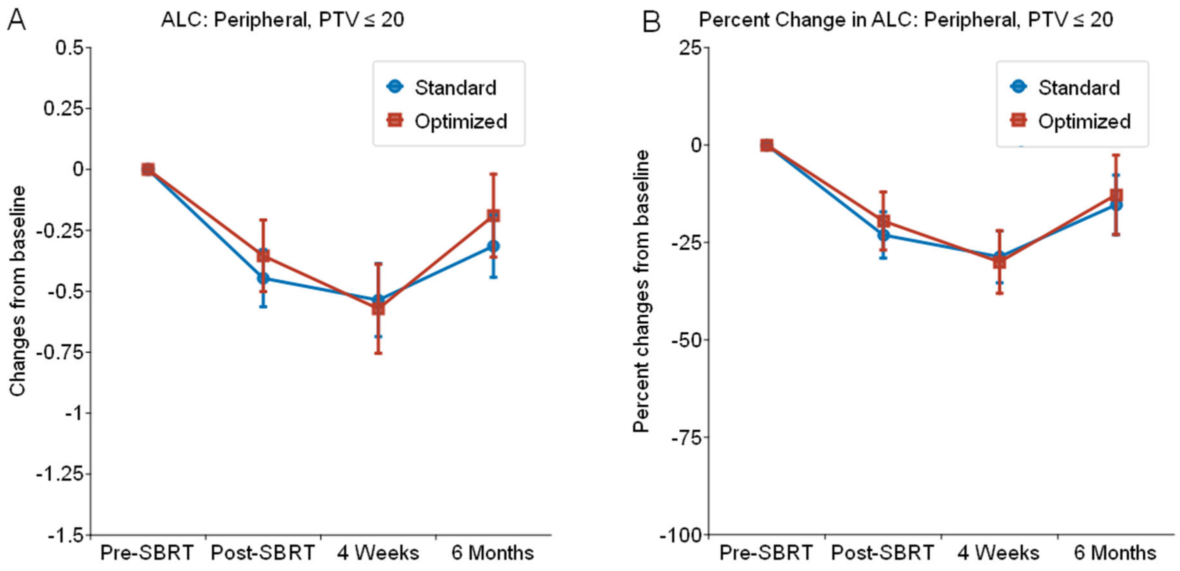


Supplementary Figure I. Average post-treatment lymphocyte counts adjusted for stratification variable, tumor location in standard and optimized arms by assessment time for peripheral tumors with PTV volume < 20 cc. Left – average absolute ALC changes from baseline. Right -Percent ALC changes as a fraction of baseline. Optimized is the experimental arm with SBRT and standard is the standard arm with SBRT.

A

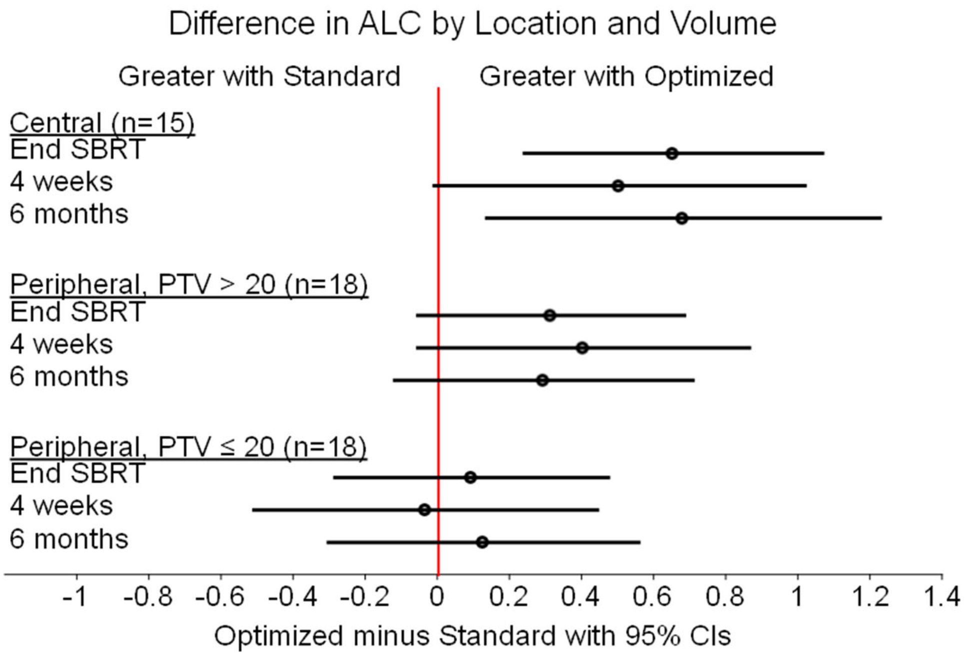


B

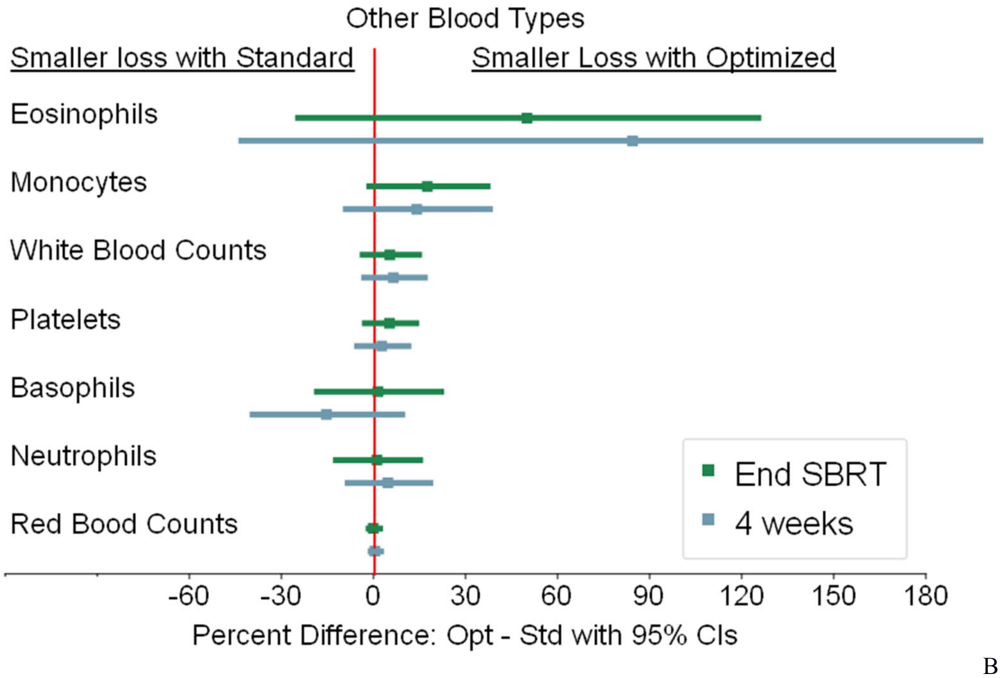


B

Supplementary Figure II: Correlation of post SBRT immune suppression with known variables. Top (A): estimated differences between optimized and standard arms in change in ALC from baseline by location and tumor volume. Bottom (B): Relative percentage reductions of different cell types atend if treatment, and at 4 weeks from SBRT.

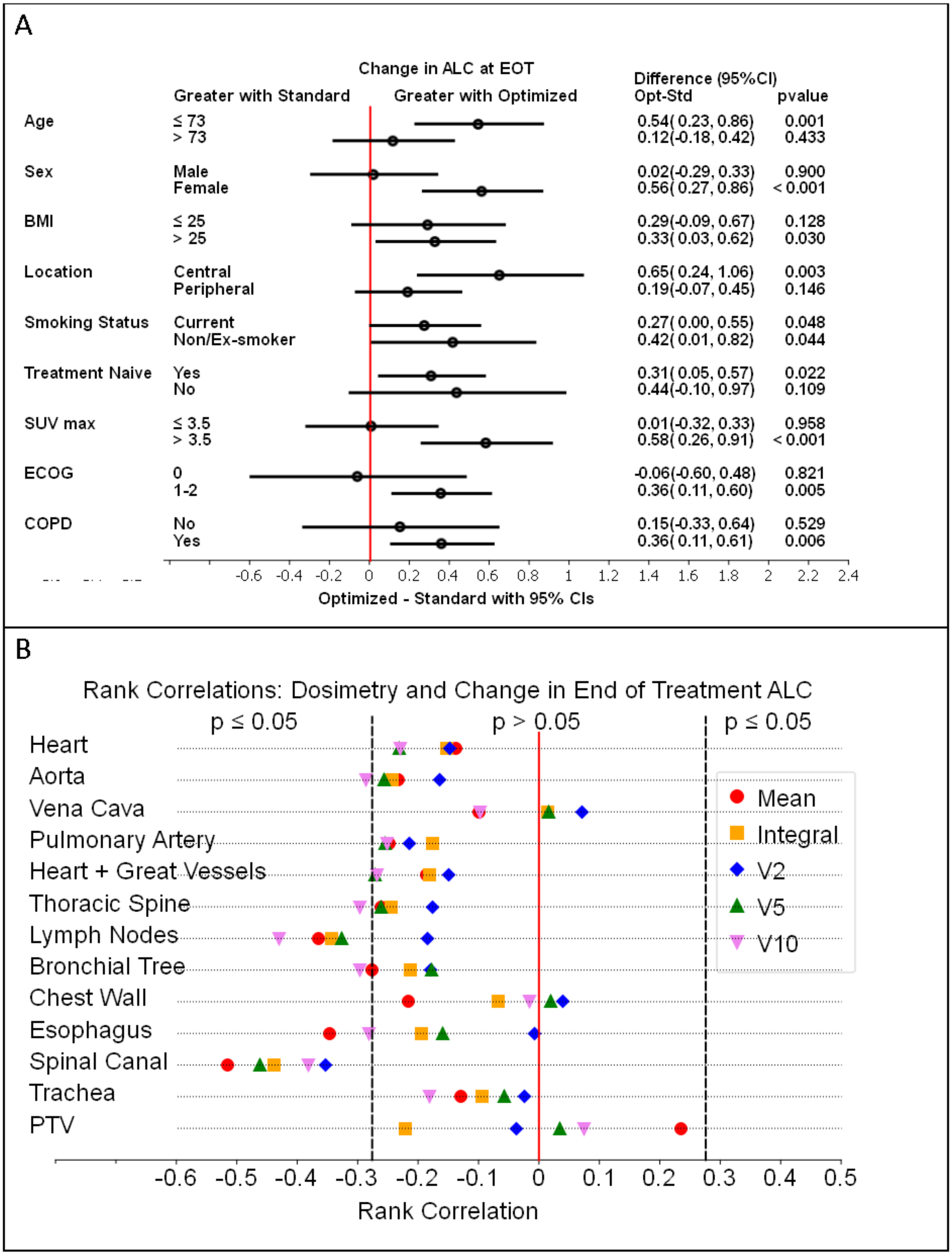


Supplementary Figure III: Correlation of post SBRT immune suppression with known variables. Top (A): estimated differences between optimized and standard arms in change in ALC at end of Tx from baseline by non-dosimetric variables. Bottom (B): estimated differences between optimized and standard arms in change in ALC at end of Tx from baseline by static organ dose volumes.

Supplementary Table I: Dosimetric Characteristics. Integral dose is the multiplication of mean dose (Gy) and total volume (cc)

| Dosimetric parameter | Optimized (n = 25) | Standard (n = 26) | Difference: Standard minus Optimized | Confidence | t-test |
|---|---|---|---|---|---|
| | Mean (range) | Mean (range) | Mean | 95% CI | p-value |
| **PTV** | | | | | |
| Mean dose | 69.5 (56.3 - 79.5) | 63.4 (54.6 - 74.8) | -6.1 | -9.5, -2.7 | 0.001 |
| Max Dose | 87.0 (66.7 - 99.9) | 71.6 (58.0 - 88.2) | -15.3 | -19.9, -10.8 | <0.001 |
| **Lungs – PTV** | | | | | |
| Volume (cc) | 4145 (2469 - 5562) | 4170 (2058 - 7414) | 25.0 | -620, 671 | 0.94 |
| Mean Dose (Gy) | 2.7 (1.9 - 5.1) | 3.0 (1.4 - 4.9) | 0.3 | 0.2, 0.8 | 0.29 |
| Integral Dose (Gy.cc) | 11210 (4781 - 17866) | 11827 (5493 - 21041) | 617.0 | -1602, 2836 | 0.58 |
| V5 (cc) | 554 (175 - 835) | 603 (236 - 1058) | 49.0 | -69, 166 | 0.41 |
| V10 (cc) | 331 (119 - 569) | 330 (150 - 632) | -1.0 | -75, 74 | 0.99 |
| V20 (cc) | 128 (59 - 247) | 130 (43 - 240) | 1.0 | -32, 34 | 0.95 |
| **External – PTV** | | | | | |
| Volume (cc) | 24923 (15753 - 31030) | 25722 (13265 - 48925) | 799.00 | -3664, 5262 | 0.72 |
| Mean Dose (Gy) | 1.2 (0.7 - 2.0) | 1.4 (0.7 - 2.9) | 0.23 | 0.0, 0.5 | 0.04 |
| Integral Dose (Gy.cc) | 29320 (19244- 49928) | 34769 (16554 - 71821) | 5449.00 | -829, 11728 | 0.09 |
| V5 (cc) | 1588 (1039 - 2982) | 1991 (886 - 4290) | 403.00 | 11, 795 | 0.04 |
| V10 (cc) | 718 (322 - 1298) | 889 (294 - 2339) | 171.00 | -49, 392 | 0.12 |
| V20 (cc) | 194 (78 - 397) | 217 (77 - 449) | 24.00 | -28, 75 | 0.36 |
| **Heart** | | | | | |
| Mean Dose (Gy) | 1.2 (0.1 - 3.4) | 1.6 (0.1 - 4.7) | 0.4 | -0.3, 1.1 | 0.25 |
| Integral Dose (Gy.cc) | 1079 (107 - 3609) | 1366 (49 - 3694) | 287 | -335, 910 | 0.36 |
| V2 (cc) | 169 (0 - 529) | 193 (0 - 737) | 24 | -82, 129 | 0.65 |
| V5 (cc) | 46 (0 - 294) | 81 (0 - 264) | 35 | -12, 82 | 0.14 |
| V10 (cc) | 6.9 (0.0 - 67.4) | 21.8 (0.0 -101.2) | 14.9 | 0.5, 29.3 | 0.04 |
| **Aorta** | | | | | |
| Mean Dose (Gy) | 1.9 (0.5 - 6.2) | 3.4 (1.0 - 9.3) | 1.5 | 0.4, 2.6 | 0.01 |
| Integral Dose (Gy.cc) | 302 (86 - 920) | 468 (146 - 1611) | 166.0 | 2, 331 | 0.05 |
| V2 (cc) | 47 (0 - 111) | 64 (13 - 156) | 17.0 | -4, 38 | 0.12 |
| V5 (cc) | 17 (0 - 66) | 33 (0 - 117) | 16.0 | 2, 30 | 0.03 |
| V10 (cc) | 3.2 (0.0 - 33.5) | 10.2 (0.0 - 75.2) | 7.0 | -0.1, 14.0 | 0.05 |
| **Vena Cava** | | | | | |
| Mean Dose (Gy) | 1.9 (0.0 - 7.2) | 3.8 (0.3 - 15.2) | 1.9 | 0.1, 3.7 | 0.04 |
| Integral Dose (Gy.cc) | 41 (0 - 206) | 65 (3 - 271) | 25.0 | -7, 56 | 0.12 |
| V2 (cc) | 6.5 (0.0 - 18.6) | 7.1 (0.0 - 17.5) | 0.6 | -2.7, 3.8 | 0.72 |
| V5 (cc) | 1.7 (0.0 - 16.0) | 4.0 (0.0 - 15.8) | 2.3 | 0.0, 4.6 | 0.05 |
| V10 (cc) | 0.6 (0.0 - 10.6) | 2.4 (0.0 - 13.6) | 1.9 | 0.1, 3.6 | 0.04 |
| **Pulmonary Artery** | | | | | |
| Mean Dose (Gy) | 2.5 (0.2 - 8.1) | 3.2 (0.0 - 18.1) | 0.7 | -1.2, 2.6 | 0.46 |

| | | | | | |
|---|---|---|---|---|---|
| Integral Dose (Gy.cc) | 163 (4 - 466) | 157 (0 - 806) | -6.0 | -108, 96 | 0.91 |
| V2 (cc) | 27.8 (0 - 86.9) | 20.2 (0 - 77.5) | -7.6 | -22.7, 7.5 | 0.32 |
| V5 (cc) | 8.6 (0 - 45.4) | 11.8 (0 - 49.5) | 3.2 | -5.4, 11.8 | 0.46 |
| V10 (cc) | 1.8 (0 - 12.5) | 3.3 (0 - 36.2) | 1.5 | -1.9, 4.9 | 0.36 |
| **Heart + Great Vessels (Aorta + Vena Cava + Pulmonary Artery)** | | | | | |
| Mean Dose (Gy) | 1.4 (0.3 - 3.7) | 1.9 (0.3 - 4.6) | 0.5 | -0.1, 1.2 | 0.12 |
| Integral Dose (Gy.cc) | 1461 (261 - 4341) | 1826 (230 - 4133) | 365.0 | -292, 1021 | 0.27 |
| V2 (cc) | 229.5 (0.0 - 587.7) | 253.8 (13.1 - 822.2) | 24.3 | -80.9, 129.6 | 0.64 |
| V5 (cc) | 67.8 (0.0 - 338.4) | 113.6 (4.6 - 300.8) | 45.8 | -5.1, 96.8 | 0.08 |
| V10 (cc) | 11.3 (0.0 - 100.5) | 32.5 (0.0 - 162.7) | 21.2 | 2.9, 39.5 | 0.02 |
| **Thoracic Spine** | | | | | |
| Mean Dose (Gy) | 1.4 (0.5 - 7.4) | 4.3 (0.7 - 9.5) | 2.9 | 1.8, 4.0 | <0.001 |
| Integral Dose (Gy.cc) | 253 (89 - 890) | 591 (136 - 2513) | 338.0 | 136, 540 | 0.002 |
| V2 (cc) | 36.9 (0.3 - 113.8) | 70.6 (31.0 - 161.1) | 33.7 | 15.6, 51.8 | <0.001 |
| V5 (cc) | 5.4 (0 - 50.2) | 40.8 (0 - 130.6) | 35.4 | 22.3, 48.5 | <0.001 |
| V10 (cc) | 1.3 (0 - 31.5) | 15.9 (0 - 87.1) | 14.6 | 6.0, 23.2 | 0.002 |
| **Lymph Node** | | | | | |
| Mean Dose (Gy) | 3.9 (1.0 - 14.2) | 6.2 (1.8 - 10.7) | 2.3 | 0.9, 3.7 | 0.002 |
| Integral Dose (Gy.cc) | 510 (117 - 1958) | 806 (221 - 1789) | 296.0 | 65, 527 | 0.01 |
| V2 (cc) | 84.5 (9.1 - 166.3) | 110.2 (32.3 - 210.1) | 25.7 | -0.3, 51.7 | 0.05 |
| V5 (cc) | 26.2 (0 - 91.3) | 62.1 (1.6 - 136.2) | 35.8 | 17.3, 54.4 | <0.001 |
| V10 (cc) | 7.3 (0 - 63.3) | 23.1 (0.0 - 72.9) | 15.8 | 5.4, 26.2 | 0.004 |
| **Bronchial Tree** | | | | | |
| Mean Dose (Gy) | 3.3 (0.2 - 17.1) | 3.2 (0.2 - 10.0) | -0.1 | -2.2, 2.0 | 0.92 |
| Integral Dose (Gy.cc) | 136 (4 - 679) | 93 (6 - 305) | -42.0 | -119, 35 | 0.27 |
| V2 (cc) | 17.1 (0.0 - 44.6) | 11.9 (0.0 - 36.5) | -5.2 | -12.7, 2.3 | 0.17 |
| V5 (cc) | 7.7 (0.0 - 36.4) | 6.3 (0.0 - 28.0) | -1.4 | -7.0, 4.2 | 0.62 |
| V10 (cc) | 3.0 (0.0 - 26.7) | 2.7 (0.0 - 14.0) | -0.2 | -3.6, 3.1 | 0.89 |
| **Chest Wall** | | | | | |
| Mean Dose (Gy) | 4.8 (2.1 - 9.3) | 5.5 (1.9 - 12.0) | 0.7 | -0.7, 2.1 | 0.32 |
| Integral Dose (Gy.cc) | 4832 (2309 - 7160) | 4816 (2033 - 9853) | -15.0 | -1008, 977 | 0.98 |
| V2 (cc) | 435.9 (200.0 - 656.0) | 406.9 (204.2 - 715.4) | -29.0 | -96.8, 38.8 | 0.39 |
| V5 (cc) | 311.6 (153.3 - 504.9) | 300.7 (121.9 - 549.7) | -10.9 | -68.0, 46.3 | 0.70 |
| V10 (cc) | 170.9 (14.5 - 321.1) | 174.8 (45.4 - 403.8) | 3.9 | -44.1, 51.9 | 0.87 |
| **Esophagus** | | | | | |
| Mean Dose (Gy) | 1.8 (0.4 - 6.8) | 2.4 (0.8 - 5.6) | 0.6 | -0.1, 1.4 | 0.11 |
| Integral Dose (Gy.cc) | 59 (21 - 266) | 71 (18 - 166) | 11.0 | -14, 36 | 0.37 |
| V2 (cc) | 8.2 (0.0 - 17.5) | 10.2 (4.0 - 25.5) | 1.9 | -0.9, 4.7 | 0.17 |
| V5 (cc) | 2.8 (0.0 - 9.4) | 4.6 (0.0 - 13.2) | 1.8 | -0.2, 3.7 | 0.07 |
| V10 (cc) | 0.9 (0.0 - 8.8) | 1.5 (0.0 - 7.4) | 0.6 | -0.6, 1.8 | 0.34 |
| **Spinal Canal** | | | | | |
| Mean Dose (Gy) | 0.9 (0.3 - 2.4) | 1.8 (0.6 - 3.4) | 0.9 | 0.5, 1.2 | <0.001 |
| Integral Dose (Gy.cc) | 46 (14 - 127) | 76 (25 - 165) | 30.0 | 10, 49 | 0.004 |

| | | | | | |
|---|---|---|---|---|---|
| V2 (cc) | 6.6 (0.0 - 21.7) | 10.6 (3.6 - 24.7) | 4.0 | 1.0, 7.1 | 0.01 |
| V5 (cc) | 1.8 (0.0 - 9.2) | 5.8 (0.0 - 12.9) | 4.0 | 2.0, 6.0 | <0.001 |
| V10 (cc) | 0.4 (0.0 - 3.0) | 1.4 (0.0 - 6.8) | 0.9 | 0.1, 1.8 | 0.04 |
| **Trachea** | | | | | |
| Mean Dose (Gy) | 1.1 (0.0 - 5.1) | 2.3 (0.1 - 8.3) | 1.2 | 0.1, 2.4 | 0.04 |
| Integral Dose (Gy.cc) | 37 (0.0 - 190) | 56.0 (1.0 - 258.0) | 19.0 | -15, 54 | 0.27 |
| V2 (cc) | 6.8 (0.0 - 33.7) | 7.7 (0.0 - 33.7) | 0.9 | -4.7, 6.4 | 0.76 |
| V5 (cc) | 1.8 (0.0 - 19.5) | 4.0 (0.0 - 26.4) | 2.2 | -0.1, 5.5 | 0.17 |
| V10 (cc) | 0.1 (0.0 - 2.1) | 1.6 (0.0 - 12.1) | 1.5 | 0.2, 2.8 | 0.02 |

Supplementary Table II: ALC summaries (raw numbers).

| Characteristic | Optimized Arm | Standard Arm | Difference: Optimized minus standard Est | 95% CI | *p* |
|---|---|---|---|---|---|
| | Mean (SD) | Mean (SD) | | | |
| Baseline ALC, (x10$^9$ cells/L) | 1.76 (0.50) | 1.87 (0.68) | -0.11 | -0.44, 0.23 | 0.53 |
| Post SBRT lymphopenia (Grade3 or higher) | 0% | 15% | -15% | -29%, -2% | 0.04 |
| Post SBRT ALC increase | 32% | 12% | 20% | -2%, 42% | 0.08 |
| **All locations (central +peripheral)** | **n=25** | **n=26** | | | |
| **End of SBRT ALC drop** | | | | | |
| Absolute [Mean (SD)] | -0.30 (0.31) | -0.61 (0.46) | 0.30 | 0.10, 0.54 | 0.01 |
| Percentage [Mean (SD)] | -15% (18%) | -30% (21%) | 15% | 4%, 26% | 0.01 |
| **At 4 weeks from SBRT initiation ALC drop** | | | | | |
| Absolute [Mean (SD)] | -0.43 (0.47) | -0.69 (0.49) | 0.26 | -0.01, 0.54 | 0.06 |
| Percentage [Mean (SD)] | -22% (24%) | -34% (18%) | 12% | 0%, 24% | 0.05 |
| **At 6 months from SBRT initiation ALC drop** | | | | | |
| Absolute [Mean (SD)] | -0.26 (0.44) | -0.51 (0.43) | 0.25 | -0.01, 0.51 | 0.06 |
| Percentage [Mean (SD)] | -15% (29%) | -24% (22%) | 10% | -5%, 25% | 0.20 |
| **Central tumor locations** | **n=9** | **n=6** | | | |
| **End of SBRT ALC drop** | | | | | |
| Absolute [Mean (SD)] | -0.20 (0.39) | -0.85 (0.55) | 0.65 | 0.05, 1.25 | 0.04 |
| Percentage [Mean (SD)] | -8% (24%) | -39%(27%) | 31% | 1%, 62% | 0.04 |
| **At 4 weeks from SBRT initiation ALC drop** | | | | | |
| Absolute [Mean (SD)] | -0.38 (0.54) | -0.89 (0.42) | 0.50 | -0.04, 1.04 | 0.07 |
| Percentage [Mean (SD)] | -18% (30%) | -43% (11%) | 26% | 1%, 50% | 0.04 |
| **At 6 months from SBRT initiation ALC drop** | | | | | |
| Absolute [Mean (SD)] | -0.32 (0.60) | -1.00 (0.22) | 0.67 | 0.14, 1.21 | 0.02 |
| Percentage [Mean (SD)] | -14% (39%) | -47% (16%) | 33% | -3%, 69% | 0.07 |
| **Peripheral, PTV>20cc** | **n=9** | **n=9** | | | |

| | | | | | |
|---|---|---|---|---|---|
| **End of SBRT ALC drop** | | | | | |
| Absolute [Mean (SD)] | -0.35 (0.23) | -0.66 (0.33) | 0.31 | 0.02, 0.60 | 0.04 |
| Percentage [Mean (SD)] | -20% (11%) | -33% (16%) | 14% | 0%, 27% | 0.05 |
| **At 4 weeks from SBRT initiation ALC drop** | | | | | |
| Absolute [Mean (SD)] | -0.37 (0.30) | -0.78 (0.47) | 0.40 | 0.01, 0.80 | 0.05 |
| Percentage [Mean (SD)] | -19% (16%) | -34% (14%) | 15% | 0%, 30% | 0.05 |
| **At 6 months from SBRT initiation ALC drop** | | | | | |
| Absolute [Mean (SD)] | -0.29 (0.36) | -0.60 (0.34) | 0.30 | -0.06, 0.67 | 0.10 |
| Percentage [Mean (SD)] | -18% (25%) | -28% (20%) | 10% | -14%, 33% | 0.39 |
| **Peripheral, PTV≤20cc** | **n=7** | **n=11** | | | |
| **End of SBRT ALC drop** | | | | | |
| Absolute [Mean (SD)] | -0.35 (0.31) | -0.45 (0.46) | 0.09 | -0.29, 0.48 | 0.62 |
| Percentage [Mean (SD)] | -20% (17%) | -23% (22%) | 4% | -16%, 23% | 0.70 |
| **At 4 weeks from SBRT initiation ALC drop** | | | | | |
| Absolute [Mean (SD)] | -0.57 (0.60) | -0.51 (0.53) | -0.06 | -0.68, 0.54 | 0.82 |
| Percentage [Mean (SD)] | -30% (26%) | -28% (23%) | -2% | -28%, 24% | 0.87 |
| **At 6 months from SBRT initiation ALC drop** | | | | | |
| Absolute [Mean (SD)] | -0.12 (0.29) | -0. 31 (0.42) | 0.20 | -0.17, 0.57 | 0.27 |
| Percentage [Mean (SD)] | -10% (17%) | -15% (20%) | 6% | -15%, 26% | 0.56 |

Supplementary Table III: ALC summaries from the statistical model estimates

| Characteristic | Optimized Arm (n = 25) | | | Standard Arm (n = 26) | | | Difference: Optimized minus standard Est | 95% CI | *p* |
|---|---|---|---|---|---|---|---|---|---|
| | Mean (SD) | 95% CI | *p* | Mean (SD) | 95% CI | *p* | | | |
| **All locations (central +peripheral)** | | | | | | | | | |
| **End of SBRT ALC drop** | | | | | | | | | |
| Absolute [Mean (SD)] | -0.31 (0.08) | -0.48, -0.15 | <0.001 | -0.65 (0.08) | -0.82, -0.48 | <0.001 | 0.33 (0.11) | 0.11, 0.56 | 0.005 |
| Percentage [Mean (SD)] | -16% (4%) | -24%, -7% | <0.001 | -31% (4%) | -39%, -22% | <0.001 | 15% (6%) | 4%, 27% | 0.01 |
| **At 4 weeks from SBRT initiation ALC drop** | | | | | | | | | |
| Absolute [Mean (SD)] | -0.45 (0.10) | -0.65, -0.25 | <0.001 | -0.73 (0.10) | -0.93, -0.53 | <0.001 | 0.28 (0.14) | 0.01, 0.56 | 0.04 |
| Percentage [Mean (SD)] | -22% (4%) | -31%, -13% | <0.001 | -34% (5%) | -43%, -25% | <0.001 | 12% (6%) | 0%, 25% | 0.05 |
| **At 6 months from SBRT initiation ALC drop** | | | | | | | | | |
| Absolute [Mean (SD)] | -0.30 (0.09) | -0.48, -0.12 | 0.002 | -0.56 (0.10) | -0.76, -0.37 | <0.001 | 0.26 (0.13) | 0.01, 0.52 | 0.04 |
| Percentage [Mean (SD)] | -16% (5%) | -27%, -5% | 0.004 | -26% (6%) | -38%, -15% | <0.001 | 10% (8%) | -5%, 26% | 0.17 |

Supplementary Table IV: Overall effect of the optimized plan versus the standard plan estimated from the model that did not include the interaction terms*

| Characteristic | Interaction p-value** | Difference: Optimized minus standard Est (SE)*** | 95% CI | *P***** |
|---|---|---|---|---|
| **All participants** | | | | |
| Absolute [Mean (SD)] | 0.76 | 0.30 (0.11) | 0.08, 0.52 | 0.007 |
| Percentage [Mean (SD)] | 0.75 | 13% (5%) | 3%, 24% | 0.01 |
| **Central tumor locations** | | | | |
| Absolute [Mean (SD)] | 0.70 | 0.64 (0.20) | 0.24, 1.04 | 0.002 |
| Percentage [Mean (SD)] | 0.79 | 30% (10%) | 10%, 49% | 0.004 |
| **Peripheral, PTV>20cc** | | | | |
| Absolute [Mean (SD)] | 0.85 | 0.31 (0.18) | -0.04, 0.67 | 0.08 |
| Percentage [Mean (SD)] | 0.93 | 14% (9%) | -4%, 21% | 0.12 |
| **Peripheral, PTV≤20cc** | | | | |
| Absolute [Mean (SD)] | 0.73 | 0.09 (0.18) | -0.27, 0.46 | 0.61 |
| Percentage [Mean (SD)] | 0.84 | 2% (9%) | -16%, 20% | 0.85 |

* For all particpants and within combinations of tumor location and tumor volume, Interaction terms were used to test whether the effect of the optimized plan versus the standard plan differed by assessment time. The results in this table did not include the interaction terms.
** This is based on the F-test for the interaction term between assessment time and treatment group. A non-significant p-value means that there is no evidence that the effects differ by assessment time.
*** This is the optimized minus the standard
**** This is the p-value for the F-test, testing whether the overall effect of optimized minus standard is zero
